# SMaRT-PCR: Sampling with Mask and Reverse Transcriptase PCR, a promising non-invasive diagnostic tool for paediatric pulmonary tuberculosis

**DOI:** 10.1101/2023.06.17.23291480

**Authors:** Ambreen Shaikh, Kalpana Sriraman, Smriti Vaswani, Ira Shah, Vishrutha Poojari, Vikas Oswal, Sushant Mane, Sakina Rajagara, Nerges Mistry

**Author notes:** **CORRESPONDENCE TO:** Kalpana Sriraman, The Foundation for Medical Research (FMR), Dr Kantilal J. Sheth Memorial Building, 84-A, R. G. Thadani Marg, Worli, Mumbai 400 018, / 24938601, Website: www.fmrindia.org.

## Abstract

**Purpose:** Diagnostic challenges in pediatric TB, like difficulties obtaining sputum, need for invasive sampling, and suboptimal sensitivity of existing detection tools, advocate for sputum-free, child-friendly, and diagnostically accurate methods. This proof-of-concept study evaluates the diagnostic potential of non-invasively **s**ampling patient bioaerosols using a mask combined with reverse transcriptase (RT)PCR (SMaRT-PCR) for pediatric TB detection.

**Methods:** In this case-control study, we recruited 51 children (30 confirmed TB and 21 without TB) aged 2-15. Exhaled bioaerosols were captured on gelatin membrane-layered N-95 masks in a 10-minute process that involved talking, coughing, and breathing. Two mask samples were consecutively collected from children with TB and tested using GeneXpert and in-house RT-PCR for *16s* and *rpoB* RNA. The control cohort underwent single mask sampling and testing with RT-PCR. Mask sampling acceptability was assessed using a questionnaire and a Likert-scale.

**Results:** The sensitivity and specificity of SMaRT-PCR for detecting *16s* and *rpoB* among 24 treatment naïve patients were 75% and 95%, respectively, comparable to GeneXpert testing in standard samples from the same patients. Mask sampling with GeneXpert had sensitivity of only 13%. Over 90% of children were comfortable with mask sampling, and > 80% were happy or very happy on the Likert scale with the procedure.

**Conclusion:** This is the first study to provide evidence for testing patient bioaerosols as a promising alternative for detecting pediatric TB. SMaRT-PCR has potential in a hub-and-spoke model, where samples collected from remote locations can be centrally tested by RT-PCR at district-level laboratories, enabling equitable access to diagnostic care.

## Introduction

Tuberculosis (TB) in children is a leading cause of death and morbidity [1]. Every year, around 1.2 million children worldwide contract TB [2], with an estimated mortality of up to 96% if untreated [3]. High-burden countries like India, Indonesia, the Philippines, Pakistan, and Nigeria account for 60.2% of pediatric TB cases [2]. Although pediatric TB is estimated to account for 12% of all incidence cases globally, only 6.9% are notified to the National Tuberculosis Programs. Thus, the gap in pediatric TB notification is estimated at 54%, rising to 69.9% in children aged 0-4 years [2].

The disparity between TB burden estimates and notifications is primarily due to difficulties diagnosing TB clinically and microbiologic confirmation of the disease in children. Children often present with nonspecific clinical and radiological findings and have difficulties expectorating sputum [4, 5]. Thus, methods such as gastric lavage, broncho-alveolar lavage, or nasopharyngeal aspiration are used to collect samples for testing. Most of these methods are invasive, distressing for children and parents, and rarely available in resource-limited settings due to the need for infrastructure and skilled personnel [6-8]. Even after obtaining the sample, due to paucibacillary TB in children, microbiological testing by smear, culture, or nucleic acid detection has a lower sensitivity [9-12]. Consequently, in a high-burden country like India, less than 25% of children are microbiologically confirmed, and a majority are treated based on clinical diagnosis [13]. Given these challenges, there has been a continued emphasis on developing child-friendly tests that use non-sputum based samples, easily obtainable in children and can be collected non-invasively, in conjunction with sensitive and accurate molecular diagnostic assays [5, 14].

In recent years, TB detection in patient respiratory aerosols has gained traction [15-18]. Exhaled or expelled aerosol samples have easy accessibility and may be used as a substitute sample, especially in patients who are unable to produce sputum [15, 17, 18]. In adult pulmonary tuberculosis (PTB) patients, our group has demonstrated that respiratory aerosols can be captured non-invasively on modified N-95 masks to detect viable TB bacteria by RT-PCR [17]. In an extended pilot analysis of 55 TB patients, aerosol Sampling with a Mask, when paired with RT-PCR (hereafter referred to as SMaRT-PCR), detected TB with 96% accuracy (unpublished data). In 10% of these patients, *Mycobacterium tuberculosis* (Mtb) bacilli could be detected in aerosols even when the patients were unable to expectorate sputum, thus indicating the potential for application in children.

The aim of this study was to test the feasibility of using mask-based sample collection for the detection of PTB in children. Mask-collected aerosols were tested for TB in children using two methods. 1. In-house-developed reverse-transcriptase quantitative PCR and 2. GeneXpert. We compared the results of mask sampling to standard-of-care sampling like gastric lavage, sputum etc. We also assessed the challenges associated with collecting mask specimens from children.

## Methods

### Participants Recruitment

Institutional Research Ethics Committees of The Foundation for Medical Research (**FMR/IREC/TB/02/2019**), Bai Jerbai Wadia Hospital for Children (BJWHC) (**IEC/BJWHC/97/2019)**, and JJ Hospital (**IEC/Pharm/RP/313/Feb-2020)** approved the study. Between August 2019 and May 2021, we recruited 51children 2-15-years of age (30 children with PTB and 21 children without TB) from three centres in Mumbai: two tertiary hospitals, - BJWHC and JJ, and a private TB clinic, Vikas Nursing Home. Written informed consent was taken from parents, and assent was taken from children aged 6-15. The TB patients included in the study either had a confirmed GeneXpert diagnosis or were clinically diagnosed based on a chest X-ray and were awaiting a GeneXpert diagnosis at the time of recruitment. Six of the 30 recruited TB patients had already started anti-TB treatment (1-3 days of treatment) by the time of study sample collection and hence were excluded from analysis for diagnosis comparison. The control group included children without a current or past history of TB for the last three years, who had a negative TST or normal chest X-ray and/or who visited the centre for alternative ailments. The TB patients had a median age of 12 (IQR 2.9-13) with 73% female, while controls had a median age of 9 (IQR 3-10) with 48% female. Supplementary Information Sheets 1 and 2 lists the clinical features of TB patients and the control group, respectively.

### SMaRT-PCR Sample Collection and Processing

We collected two mask samples from each TB patient (for RT-PCR and GeneXpert/Ultra) and one from control participant (for RT-PCR). For sampling, the participants wore a modified flat-fold (3M 9210) or cup-type (3M 8110s) N-95 mask with a 37-mm diameter gelatin membrane (Sartorius, Gottingen, Germany) attached to the inner surface of the mask. The participant performed directed tasks for ten minutes, including two rounds of talking, reading, or reciting for about 3 minutes based on the child’s preference, one minute of coughing, and one minute of tidal breathing. After 10 minutes, the membrane was removed with forceps and transferred to a collection cup containing 3 ml of RNAzol™ (Sigma-Aldrich, MO, USA) for RT-PCR testing or 4 ml of PBS for GeneXpert/Ultra (Cepheid, USA) testing. Mask-sampling acceptance among children was assessed using a questionnaire and a Likert scale survey tool (Supplementary Information Sheet 3).

All RT-PCR assays were performed at FMR and GeneXpert at BJWHC. For RT-PCR testing, the total RNA was extracted from RNAzol™ containing the dissolved gelatin membrane, purified using an RNeasy Micro RNA Isolation Kit (Qiagen, Hilden, Germany), treated with DNase I (New England Biolabs, Massachusetts, USA) and reverse transcribed using the Iscript cDNA Synthesis Kit (Bio-Rad, California, USA) as described earlier [17]. For the detection of Mtb, we evaluated the amplification (Ct values) and product-specific melt peaks of Mtb-specific genes *16S* and *rpoB* using SYBR green dye in the CFX96 Touch Real-time PCR detection system (Bio-Rad). The samples were denoted as positive if the *16S* and/or *rpoB* Ct values were < 40 and had a product-specific melt curve. For samples with a Ct value > 35 for either of the genes, the amplification was confirmed by running the qPCR product on a 2% agarose gel. All analysis for the diagnostic performance of SMaRT-PCR for TB was considered for two outcomes (positive for *16S* **OR** *rpoB*, positive for both *16S* **AND** *rpoB*)

## Results

### Comparison of SMaRT-PCR with GeneXpert in standard samples and mask samples for TB detection

For the 24 treatment-naïve TB patients, GeneXpert was applied for diagnosis in sputum (14) or induced sputum (1), or gastric lavage (GL) (8), or nasal aspirate (1) samples. In these samples, GeneXpert detected TB in 17/24 (71%) patients. In the seven GeneXpert-negative patients, the TB diagnosis was made by smear AFB positivity (1), clinical presentation and chest X-ray (6).

In comparison, SMaRT-PCR detected TB in 23/24 (96%) patients when the condition for detection was the presence of either the 16S or rpoB gene and in 18/24 (75%) when the condition was both genes (Supp Table S1). In the concomitantly collected second mask sample tested by GeneXpert, TB could be detected only in 3/24 (13%) patients. All three patients had only trace quantities detected, indicating that the GeneXpert sensitivity in mask samples was very low.

### SMaRT-PCR of control patients

Among the 21 control patients with no known TB, one gene (*16S or rpoB*) could be detected in 4 patients (Supp Table S2 Control Patients 2, 3,11, and 17), and both genes (*16S* and *rpoB*) could be detected in 2 patients (Supp Table S2 Control Patients 2 and 17). Control patients in whom Mtb-specific genes were detected were followed up for a potential TB diagnosis. Patient 2 underwent a chest HRCT scan, which showed mediastinal adenopathy with bilateral axillary adenopathy. Since the patient’s three siblings had EPTB, with one sibling on TB treatment, patient 2 was clinically diagnosed to have TB and was initiated on TB treatment. Control patient 17 underwent an HRCT scan of the chest, GeneXpert, and culture test from the GL sample. Both GL-GeneXpert and culture were negative for TB; however, the HRCT showed fibrotic changes in the right lower lobe with calcification, and the mediastinal lymph node showed signs of post-infection sequelae. Thus, the treating physician decided not to initiate TB treatment. Since control patient 2 was clinically diagnosed to have TB retrospectively, this patient was excluded from specificity analysis for SMaRT-PCR. The other control patients (patient IDs 3,11,17) with at least one gene detected were considered false positives by SMaRT-PCR.

### Sensitivity and Specificity of SMaRT-PCR

The SMaRT-PCR’s sensitivity and specificity were calculated based on 24 treatment-naïve TB patients and 20 controls and at two conditions for detection-a) positive, if either *16S* OR *rpoB* was detected; b) positive, only if both *16S* AND *rpoB* were detected (Table-1). If 16S OR rpoB was present, SMaRT-PCR had 96% sensitivity and 85% specificity. If both *16S* AND *rpoB* were considered, the sensitivity decreased to 75% while the specificity increased to 95%. GeneXpert had a sensitivity of 71% in our sample set and was reported to have 98% specificity in such samples [9].

**Table-1:**
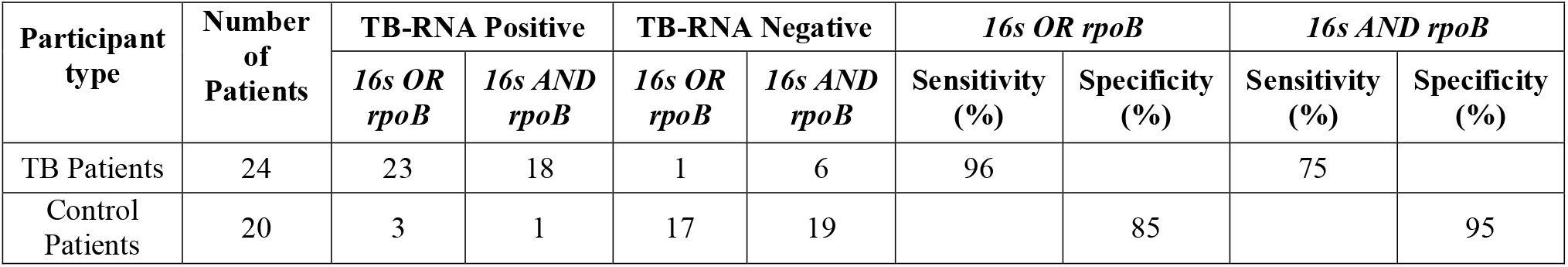
Sensitivity and Specificity of SMaRT-PCR.

| Participant type | Number of Patients | TB-RNA Positive |  | TB-RNA Negative |  | *16s OR rpoB* |  | *16s AND rpoB* |  |
| --- | --- | --- | --- | --- | --- | --- | --- | --- | --- |
|  |  | *16s OR rpoB* | *16s AND rpoB* | *16s OR rpoB* | *16s AND rpoB* | Sensitivity (%) | Specificity (%) | Sensitivity (%) | Specificity (%) |
| TB Patients | 24 | 23 | 18 | 1 | 6 | 96 |  | 75 |  |
| Control Patients | 20 | 3 | 1 | 17 | 19 |  | 85 |  | 95 |

### SMaRT-PCR of TB patients under treatment

Six of the 30 TB patients recruited (Supp. Table S1; patients #9-13 and 17) had received anti-TB treatment for 1-3 days by the time of study sample collection. All these patients were TB positive by GeneXpert in GL (patients 9-13) or BAL (patient 17) samples before treatment initiation. The TB positivity by SMaRT-PCR was 3/6 for *16S* AND *rpoB* and 4/6 for *16S* OR *rpoB*. The results indicate that SMaRT-PCR’s ability to detect TB may decrease if the patients are initiated on appropriate anti-TB treatment.

### Study participants of interest

#### TB patient 20

This participant had 10 days of cough and weight loss for 90 days. The chest X-ray showed infiltrates in the left middle and lower lungs, suggesting primary progressive TB. The patient’s consecutive sputum and nasal aspirate sample tested negative for TB by GeneXpert. However, the treating physician diagnosed TB based on clinical and X-ray presentation and initiated her on TB treatment. Interestingly, both mask samples collected from this patient were positive for TB by RT-PCR and GeneXpert, affirming the clinical diagnosis made without knowledge of mask sample results.

#### Control patient 2

This participant had no symptoms but underwent a chest X-ray as one of the participant’s siblings was being treated for EPTB at the time of approaching the physician, and two other siblings had completed their treatment for EPTB. His physical and chest X-ray examination was normal. Since the child was asymptomatic with a normal chest X-ray, the physician ruled out TB and referred him to us as a control patient. However, SMaRT-PCR was positive for both *16S* and *rpoB*. Following which, as mentioned before, the patient underwent chest-HRCT, which showed mediastinal adenopathy (1 node of 1.5×1.2 cm) and bilateral axillary adenopathy (2 enlarged nodes of 1.1×1 cm and 1×0.8 cm) and no involvement of lung parenchyma. Since the patient had three siblings who had EPTB in the recent past and an aunt with a history of PTB 4 years prior to the time of recruitment, the physician concluded that the child had TB and initiated him on TB treatment. After almost 3 months of anti-TB treatment, repeat HRCT showed only bilateral axillary adenopathy with multiple lymph nodes of 1.5× 1.6 cm. After 8 months of anti-TB treatment, the ultra-sonography of the axilla showed residual non-necrotic lymph nodes in the right axilla (1.3 ×0.7cm; 0.5×0.5cm) and two necrotic lymph nodes in the left axilla (2 ×0.8cm; 0.6×0.2 cm), suggesting a positive response to treatment.

### Acceptability of sampling with mask

The sample collectors noted the acceptability of the mask sampling method among the children (Table-2). Over 90% of children performed the directed tasks and were comfortable with the sampling procedure. A small proportion of TB patients (4/30) struggled to perform the tasks and/or complete the 10-minute sampling time, either due to tachypnea or difficulty comprehending the task requirement. Only 67% of the children expressed a willingness to repeat the test, indicating there may be some challenges to repeating tests if necessary.

**Table 2:**
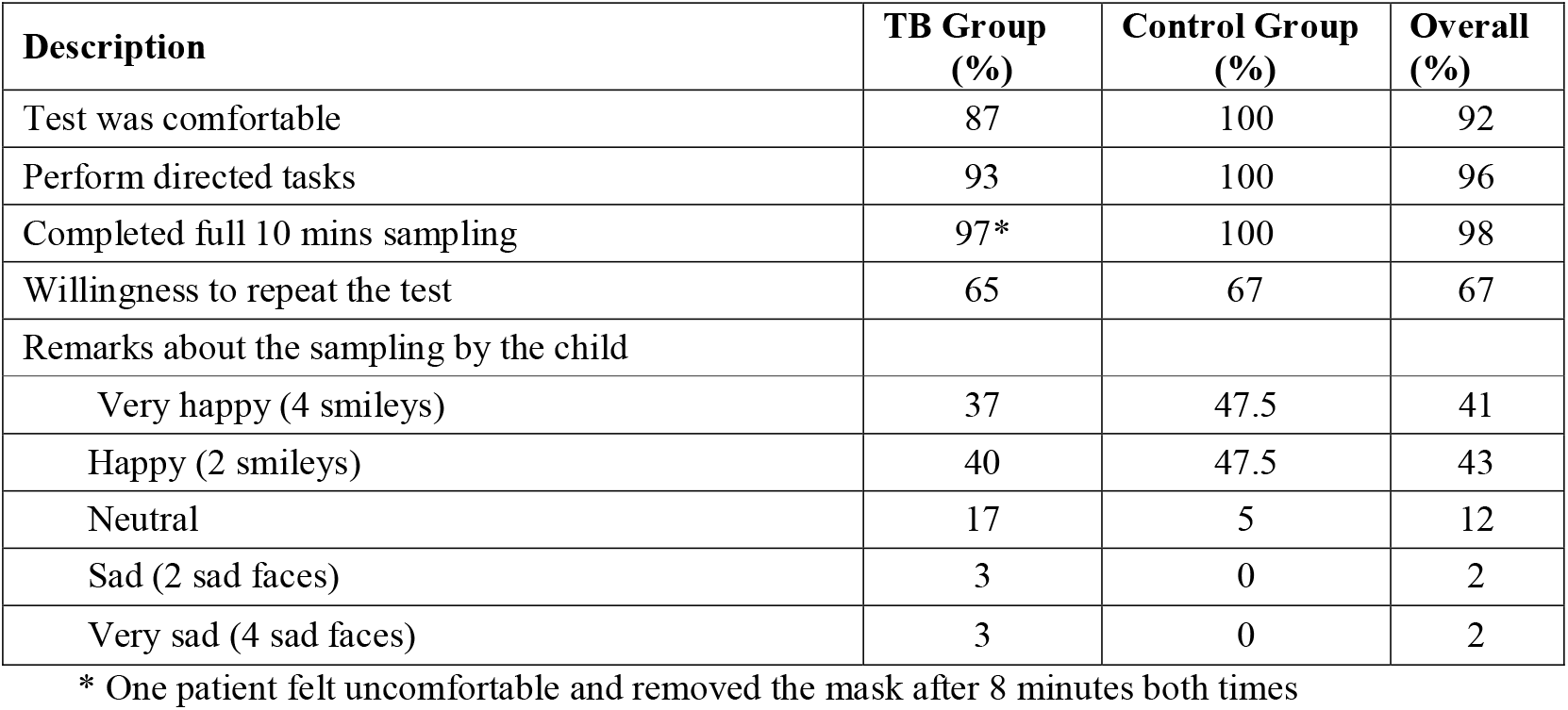
Patient Acceptability of Sample Collection.

## Discussion

Microbiological confirmation of pulmonary TB in children often requires invasive sampling, and TB detection in these samples is suboptimal. The current study investigated the feasibility of diagnosing TB in children by capturing infectious aerosols exhaled/emitted by them while talking, coughing, and breathing using an adapted N95 mask. The sampling method was combined with the standard GeneXpert testing as well as an in-house RT-PCR to detect TB-specific RNA. Here, we showed for the first time that mask sampling can be a promising non-invasive alternative for pediatric TB detection. When paired with an in-house RT-PCR, the method detected TB with 75% sensitivity and 95% specificity in GeneXpert-positive or clinically diagnosed paediatric TB patients, comparable to the standard diagnosis method. Interestingly, the method detected TB in one patient where standard methods failed and in one control patient with undetected TB in the Lymph node. Other non-invasive specimens for detecting pediatric TB, like urine, stool, or oral swabs, have reported sensitivity and specificity between 37-63% and 85-98%, respectively [19-22]. The SMaRT-PCR performed better than these methods in sensitivity and had comparable specificity. However, authors concur that the current SMaRT-PCR results represent a limited sample size and that larger field trials in presumptive patients are imperative to evaluate the method’s true diagnostic accuracy.

This study found that mask aerosol sampling with GeneXpert had low sensitivity (13%). Similar low sensitivity was also observed when oral swab sampling (21%, [23]) and tongue swab sampling (26%, [24]) were combined with GeneXpert for detecting pediatric TB. One possible explanation could be that GeneXpert is not optimized for detecting nontraditional samples like mask-captured aerosols or oral swabs. Our in-house RT-PCR outperformed GeneXpert in the mask specimen sensitivity (75% vs 13%). The increased sensitivity could be attributed to techniques like sample concentration [25, 26], precise RNA isolation, and longer RT-PCR amplification runs of up to 40 cycles [24, 27, 28], all known to improve sensitivity. Another possible reason could be the detection of abundantly expressed RNA by RT-PCR vs fewer copies of DNA detected in GeneXpert. SMaRT-PCR detects 16S rRNA, which has higher copy numbers compared to IS element copies in the GeneXpert-detected Mtb DNA [29]. All these factors suggest that molecular GeneXpert testing of atypical samples requires more optimization research.

The most significant feature of the SMaRT-PCR procedure is the non-invasive sample collection, which was well received by children who underwent mask sampling. Collecting aerosols on the mask is simple, quick, and painless, with low risk to the sample collector when an age-appropriate N95 mask is worn. Furthermore, the SMaRT-PCR assay has the potential to be used in a hub-and-spoke model, with its non-invasive and easy sample collection that can be decentralized to the primary levels of the health system, and sample processing can be done at the district-level lab equipped with a real-time PCR machine. Sample collection decentralization will also enable SMaRT-PCR to reach more children in inaccessible areas, thus enabling equitable access. Although RT-PCR is a moderately complex technology, the capacity to use this technology has been strengthened globally during the COVID-19 pandemic. Alternatively, technologies like GeneXpert or TrueNat can also be adapted for performing RT-PCR, as shown for COVID-19 RT-PCR [30, 31]. We envision the SMaRT-PCR integrating with these up-skilled and strengthened facilities for future programmatic use.

The WHO target product profile (TPP) for non-sputum diagnostic tests for PTB in children requires sensitivity > 66% and 98% specificity for microbiologically confirmed TB [32]. Within our tested cohort, the SMaRT-PCR method met the requisite sensitivity but had a 95% specificity. TPP development and increased advocacy have led to the creation of several novel tests [33]. However, none of the available non-sputum samples like urine, stool, or oral swabs meets the complete TPP requirements [19-22]. Though no single test has the required sensitivity or specificity, larger studies combining two or more samples may provide better answers on how to integrate these diagnostic approaches in a complementary manner in an algorithm to reach the WHO TPP. A good example would be the proven 8-20% increase in GeneXpert’s diagnostic yield by testing an additional specimen [34].

Our proof-of-concept pilot study had certain limitations; the testing was done in a small cohort of confirmed TB patients, and thus we could not stratify patients by factors such as age or nutritional or HIV status, all of which could influence the SMaRT-PCR’s diagnostic accuracy. Treatment follow-up studies that could classify patients as unconfirmed or unlikely TB based on symptom resolution were not undertaken. Such long-term follow-up studies are necessary for proper diagnostic accuracy estimates[1]. This study was limited to mask sampling in tertiary hospitals within an hour of the sample processing lab, so it could not assess the impact of long-term sample storage and transport on diagnostic yield. Based on our learnings from the proof-of-concept study, we have refined and improved the SMaRT-PCR to enable remote sample collection with a simplified RNA isolation protocol and a probe-based, multiplex RT-PCR assay that can simultaneously detect multiple TB-specific genes from a small sample volume and determine TB drug resistance. This refined SMaRT-PCR awaits diagnostic performance evaluation in larger field studies.

## Conclusion

Overall, our pilot investigation provides promising data on the potential application of a non-invasive SMaRT-PCR for detecting viable Mtb in aerosols exhaled or expelled by children. The use of a SMaRT-PCR workflow for children could aid in early TB confirmation, therefore possibly improving TB detection rates and paving the way for better treatment outcomes. Towards achieving this impact, in the short term, we propose to scale the refined workflow for multisite testing in a larger sample size with longitudinal follow-up throughout treatment to better evaluate diagnostic accuracy in presumptive pulmonary pediatric TB populations. The multisite studies would aid in: 1. comparing the performance of the SMaRT-PCR to existing molecular methods and the gold standard of TB culture. 2. examining any correlation between SMaRT-PCR positivity and patient demographics or disease features 3. determining if SMaRT-PCR can be used as a standalone rule-in test or applied in complementary or cafeteria approach more suited for a specific population at a specific stage of TB.

## Data Availability

The data supporting this study's findings are available from the corresponding author upon reasonable request.

## Acknowledgements

This research study was supported through private donation to FMR from Mr. Nadir Godrej (Indian Industrialist) and FMR institutional funds. We thank Dr Varinder Singh, Director, Professor of Pediatrics, Lady Hardinge Medical College and Assoc Kalawati Saran Children’s Hospital, New Delhi, India, for his support, inputs and initial encouragement to test it in children. We thank India Health Fund, which funded the testing initially in adults and facilitated association with central TB decision for the study. We thank Dr Raghuram Rao, Central TB division, for facilitating JJ Hospital as a site for the study. We sincerely appreciate the contribution of Mr Nilesh Shahasane, the field assistant at FMR, who collected and transported samples. Finally, we would like to thank all the participants of this study and their parents for their cooperation, patience, and support, without which this study would not have been possible.

## Statements and Declarations

### Author Contributions (Contributor Roles Taxonomy)

Conceptualization: KS, NM. Data curation: AS, KS, SV, VK. Formal Analysis: KS, AS. Funding acquisition: KS, NM. Investigation: AS, SV, VK, SR. Methodology: AS, KS, IS. Project administration: KS, AS. Resources: IS, VO, SM. Supervision: NM, IS, VO, SM. Writing – original draft: AS KS. Writing – review & editing: All authors. IS, VO and SM are Senior Clinical Collaborators who have contributed substantially and equally to the study.

### Ethics

The study was undertaken after the approval of the Institutional Research Ethics Committees of The Foundation for Medical Research (FMR/IREC/TB/02/2019), Bai Jerbai Wadia Hospital for Children (IEC/BJWHC/97/2019), and JJ Hospital (IEC/Pharm/RP/313/Feb-2020)

### Consent

The authors affirm that parents of all research participants provided informed written consent to participate and publish

## References

[1] Olbrich L, Nliwasa M, Sabi I, Ntinginya NE, Khosa C, Banze D, Corbett EL, Semphere R, Verghese VP, Michael JS, Graham SM, Egere U, Schaaf HS, Morrison J, McHugh TD, Song R, Nabeta P, Trollip A, Geldmacher C, Hoelscher M, Zar HJ, Heinrich N (2023) Rapid and Accurate Diagnosis of Pediatric Tuberculosis Disease: A Diagnostic Accuracy Study for Pediatric Tuberculosis. Pediatr Infect Dis J 42 (5):353–360

[2] WHO (2022) Global Tuberculosis Report.

[3] Dodd PJ, Yuen CM, Sismanidis C, Seddon JA, Jenkins HE (2017) The global burden of tuberculosis mortality in children: a mathematical modelling study. Lancet Glob Health 5 (9):e898–e906

[4] Edwards D, Kitetele F, Van Rie A (2007) Agreement between clinical scoring systems used for the diagnosis of pediatric tuberculosis in the HIV era. The International Journal of Tuberculosis and Lung Disease 11 (3):263–269

[5] Organization WH (2018) Roadmap towards ending TB in children and adolescents.

[6] 2020 GWHO (2020) Global tuberculosis report 2020: executive summary.

[7] Togun T, Kampmann B, Stoker N, Lipman M (2020) Anticipating the impact of the COVID-19 pandemic on TB patients and TB control programmes. Annals of Clinical Microbiology and Antimicrobials.

[8] Zar HJ, Hanslo D, Apolles P, Swingler G, Hussey G (2005) Induced sputum versus gastric lavage for microbiological confirmation of pulmonary tuberculosis in infants and young children: a prospective study. The Lancet 365 (9454):130–134

[9] Detjen AK, DiNardo AR, Leyden J, Steingart KR, Menzies D, Schiller I, Dendukuri N, Mandalakas AM (2015) Xpert MTB/RIF assay for the diagnosis of pulmonary tuberculosis in children: a systematic review and metaanalysis. Lancet Respir Med 3 (6):451–461

[10] Kunkel A, Abel Zur Wiesch P, Nathavitharana RR, Marx FM, Jenkins HE, Cohen T (2016) Smear positivity in paediatric and adult tuberculosis: systematic review and meta-analysis. BMC Infect Dis 16:282

[11] Perez-Velez CM, Marais BJ (2012) Tuberculosis in children. New England Journal of Medicine 367 (4):348–361

[12] Sharma G, Malhotra B, John PJ, Gautam S, Bhargava S (2022) Evaluation of GeneXpert and liquid culture for detection of Mycobacterium tuberculosis in pediatric patients. Indian Journal of Medical Microbiology 40 (4):547–551

[13] Central TB Division M (2022) India TB Report.

[14] Denkinger CM, Kik SV, Cirillo DM, Casenghi M, Shinnick T, Weyer K, Gilpin C, Boehme CC, Schito M, Kimerling M (2015) Defining the needs for next generation assays for tuberculosis. The Journal of infectious diseases 211 (suppl_2): S29–S38

[15] Fennelly KP, Acuna-Villaorduna C, Jones-Lopez E, Lindsley WG, Milton DK (2020) Microbial Aerosols: New Diagnostic Specimens for Pulmonary Infections. Chest 157 (3):540–546

[16] Jones-López EC, Acuña-Villaorduña C, Ssebidandi M, Gaeddert M, Kubiak RW, Ayakaka I, White LF, Joloba M, Okwera A, Fennelly KP (2016) Cough aerosols of Mycobacterium tuberculosis in the prediction of incident tuberculosis disease in household contacts. Clinical Infectious Diseases 63 (1):10–20

[17] Shaikh A, Sriraman K, Vaswani S, Oswal V, Mistry N (2019) Detection of Mycobacterium tuberculosis RNA in bioaerosols from pulmonary tuberculosis patients. Int J Infect Dis 86:5–11

[18] Williams CM, Cheah ES, Malkin J, Patel H, Otu J, Mlaga K, Sutherland JS, Antonio M, Perera N, Woltmann G, Haldar P, Garton NJ, Barer MR (2014) Face mask sampling for the detection of Mycobacterium tuberculosis in expelled aerosols. PLoS One 9 (8):e104921

[19] Gebre M, Cameron LH, Tadesse G, Woldeamanuel Y, Wassie L (2021) Variable Diagnostic Performance of Stool Xpert in Pediatric Tuberculosis: A Systematic Review and Meta-analysis. Open Forum Infect Dis 8 (8):ofaa627

[20] Nicol MP, Allen V, Workman L, Isaacs W, Munro J, Pienaar S, Black F, Adonis L, Zemanay W, Ghebrekristos Y, Zar HJ (2014) Urine lipoarabinomannan testing for diagnosis of pulmonary tuberculosis in children: a prospective study. Lancet Glob Health 2 (5):e278–284

[21] Nicol MP, Wood RC, Workman L, Prins M, Whitman C, Ghebrekristos Y, Mbhele S, Olson A, Jones-Engel LE, Zar HJ, Cangelosi GA (2019) Microbiological diagnosis of pulmonary tuberculosis in children by oral swab polymerase chain reaction. Scientific Reports 9 (1):10789

[22] Wang Y, He Y, Wang L, Zhang Y-A, Wang M-S (2023) Diagnostic Yield of Nucleic Acid Amplification Tests in Oral Samples for Pulmonary Tuberculosis: A Systematic Review and Meta-analysis. Open Forum Infectious Diseases 10 (3)

[23] Cox H, Workman L, Bateman L, Franckling-Smith Z, Prins M, Luiz J, Van Heerden J, Ah Tow Edries L, Africa S, Allen V, Baard C, Zemanay W, Nicol MP, Zar HJ (2022) Oral Swab Specimens Tested With Xpert MTB/RIF Ultra Assay for Diagnosis of Pulmonary Tuberculosis in Children: A Diagnostic Accuracy Study. Clinical Infectious Diseases 75 (12):2145–2152

[24] Flores JA, Calderón R, Mesman AW, Soto M, Coit J, Aliaga J, Mendoza M, Leon SR, Konda K, Mestanza FM, Mendoza CJ, Lecca L, Murray MB, Holmberg RC, Pollock NR, Franke MF (2020) Detection of Mycobacterium Tuberculosis DNA in Buccal Swab Samples from Children in Lima, Peru. Pediatr Infect Dis J 39 (11):e376–e380

[25] Song Y, Ma Y, Liu R, Shang Y, Ma L, Huo F, Li Y, Shu W, Wang Y, Gao M, Pang Y (2021) Diagnostic Yield of Oral Swab Testing by TB-LAMP for Diagnosis of Pulmonary Tuberculosis. Infect Drug Resist 14:89–95

[26] Steingart KR, Ng V, Henry M, Hopewell PC, Ramsay A, Cunningham J, Urbanczik R, Perkins MD, Aziz MA, Pai M (2006) Sputum processing methods to improve the sensitivity of smear microscopy for tuberculosis: a systematic review. Lancet Infect Dis 6 (10):664–674

[27] LaCourse SM, Pavlinac PB, Cranmer LM, Njuguna IN, Mugo C, Gatimu J, Stern J, Walson JL, Maleche-Obimbo E, Oyugi J, Wamalwa D, John-Stewart G (2018) Stool Xpert MTB/RIF and urine lipoarabinomannan for the diagnosis of tuberculosis in hospitalized HIV-infected children. Aids 32 (1):69–78

[28] Luabeya AK, Wood RC, Shenje J, Filander E, Ontong C, Mabwe S, Africa H, Nguyen FK, Olson A, Weigel KM, Jones-Engel L, Hatherill M, Cangelosi GA (2019) Non-invasive Detection of Tuberculosis by Oral Swab Analysis. J Clin Microbiol 57 (3)

[29] Backstedt BT, Buyuktanir O, Lindow J, Wunder Jr EA, Reis MG, Usmani-Brown S, Ledizet M, Ko A, Pal U (2015) Efficient detection of pathogenic leptospires using 16S ribosomal RNA. PLoS One 10 (6):e0128913

[30] Rakotosamimanana N, Randrianirina F, Randremanana R, Raherison MS, Rasolofo V, Solofomalala GD, Spiegel A, Heraud J-M (2020) GeneXpert for the diagnosis of COVID-19 in LMICs. The Lancet global health 8 (12):e1457–e1458

[31] Sadhna S, Hawaldar R (2020) Evaluation of Truenat RT PCR for diagnosis of SARS CoV2 infection-An observational study. Indian J Microbiol Res 7 (3):265–269

[32] World Health O (2014) High priority target product profiles for new tuberculosis diagnostics: report of a consensus meeting, 28–29 April 2014, Geneva, Switzerland. World Health Organization, Geneva

[33] Nicol MP, Zar HJ (2020) Advances in the diagnosis of pulmonary tuberculosis in children. Paediatr Respir Rev 36:52–56

[34] Zar HJ, Workman LJ, Prins M, Bateman LJ, Mbhele SP, Whitman CB, Denkinger CM, Nicol MP (2019) Tuberculosis diagnosis in children using Xpert Ultra on different respiratory specimens. American journal of respiratory and critical care medicine 200 (12):1531–1538

